# Axonal degeneration serum markers and temporal lobe atrophy in Alzheimer’s dementia continuum: a longitudinal study of plasma neurofilament light and tensor-based morphometry

**DOI:** 10.1101/2024.03.21.24304687

**Authors:** Amirhossein Khodadadi, Nikan Amirkhani, Zahra Nouri, Tanin Adlparvar, Sanaz Eskandari, Rezvan Barzgar, Pouya Sojoudi, Mahsa Mayeli

**Affiliations:** Mashhad University of medical sciences; Tehran University of medical sciences; University of Tehran; Tabriz Islamic Azad University; Sirjan Payame Noor University; Technical University of Munich; University of Maryland

**Keywords:** Alzheimer’s disease, mild cognitive impairment, biomarker, MRI

## Abstract

The plasma neurofilament light chain (NfL), an axonal cytoskeleton protein, increases in Alzheimer’s disease and was therefore proposed as a blood-based biomarker of the disease. Tensor-based morphometry (TBM) is an MR based modality that identifies local volume changes in the brain. Herein, we aimed to investigate whether plasma NfL measures can predict TBM findings derived from temporal lobe of brain in a one-year follow-up and which biomarker can predict cognitive function. A total of 480 participants with Alzheimer’s disease (AD), mild cognitive impairment (MCI), and normal cognition (CN) were found eligible for inclusion from The Alzheimer’s Disease Neuroimaging Initiative (ADNI) database. There was a significant negative association between plasma NfL and TBM only when all subjects were pooled together at baseline (β = -0.139, *P=* 0.004). After one-year follow-up, 30 subjects with MCI converted to AD (MCI-AD) and others remained unchanged (CN, MCI, AD). Plasma NfL levels elevated significantly only in MCI group after one year (*P*<0.001). We found a significant reduction in TBM measurements at first-year compared to baseline in all groups (*P*<0.001 for all groups). Additionally, TBM average change rate was significantly higher in MCI-AD and AD groups (*P*<0.001 for both); however, plasma NfL average change rate was not significantly different between groups. TBM was significantly correlated with MMSE, MoCA, ADAS-11 and ADAS-13 scores in both MCI and AD patients at baseline and after one year, whereas plasma NfL was not. Overall, our findings indicate that plasma NfL is not reliably associated with TBM, and is less effective and sensitive than TBM in predicting dementia progression and cognitive performance. Hence, TBM reduction is not reflected in plasma NfL increment after one year follow-up.

## 1. Introduction

Alzheimer’s disease (AD) is known to be the most common cause of dementia [1] and is characterized by the deposition of beta-amyloid plaques and neurofibrillary tangles in the brain [2]. The prevalence of AD is closely linked with increasing age, and the elderly (>65 years) population has been projected to increase from 7% in 2000 to approximately 12% in 2030 worldwide [3]. Therefore, this trend establishes an expectation for a corresponding rise in AD prevalence. AD is usually diagnosed at the late stages when the disease has already progressed and disease-modifying therapies are no longer effective [4]. Early detection of prodromal and preclinical AD is therefore essential [5]. This necessitates the identification of robust biomarkers, including cerebrospinal fluid (CSF), plasma, and neuroimaging markers to enable early diagnosis.

CSF biomarkers have been extensively studied as measures of early AD detection and have had high diagnostic accuracy, while plasma biomarkers have had less success. However, due to the less invasive nature of plasma samples compared to CSF, plasma-based markers have a higher chance of patient approval as screening measures. Neurofilament light chain (NfL) is one such proposed plasma biomarker for neurodegeneration in dementia [6].

Low levels of NfL are constantly released from neurons into the extracellular space and ultimately reach the cerebrospinal fluid (CSF) and blood [6]. However, axonal damage leads to a sharp increase in NfL release, and consequently, increased plasma NfL concentrations are reported in neurodegenerative and neuroinflammatory diseases [7]. Increased plasma NfL has been suggested to be associated with early AD diagnosis 10 to 20 years in advance of clinical symptoms [8].

The degenerative pathology of AD can also be observed as local volume changes in magnetic resonance (MR) images of the brain. Tensor-based morphometry (TBM) is an image analysis technique that identifies these regional differences from the gradients of the nonlinear deformation fields that align images to a standard anatomical atlas [9]. Also, deep grey matter structural abnormalities in patients with MCI and AD and how they relate to cognition can be assessed using TBM [10]. TBM-derived measures of brain atrophy can be used as an imaging biomarker for AD and have been observed to correlate well with clinical measures of cognitive deterioration, including CDR, MMSE, and memory tests [11]. The alignment of plasma and imaging biomarkers of AD can help with the early detection of patients without the need for the relatively invasive techniques of CSF collection. However, studying the accordance between these markers has not been undertaken extensively. In this research, we assessed the correlation between the imaging biomarkers resulting from TBM analysis of the brain’s temporal lobe and plasma NfL concentration across the spectrum of dementia. Specifically, we aim to answer whether TBM index decrement is reflected with plasma NfL increment and which biomarker is reliably associated with disease progression and cognitive performance.

## 2. Methods and materials

### 2.1. Data collection

The Alzheimer’s Disease Neuroimaging Initiative (ADNI) Database was used to acquire data for this study. ADNI is a longitudinal multicenter study aimed at developing different clinical, imaging, genetic, and biochemical biomarkers to detect Alzheimer’s disease at the very early stage. ADNI started in 2004 under the leadership of Dr. Michael W. Weiner, the principal investigator. It was funded as a private-public partnership. To date, three different phases of ADNI have been undertaken. For further information on this aspect, please refer to https://adni.loni.usc.edu/.

### 2.2. Participants

A number of 157 cognitively normal (CN), 278 mild cognitive impairment (MCI), and 45 AD subjects were found eligible for inclusion at baseline. The number of subjects per diagnostic group changed after one-year follow-up (HC=154, MCI=253, AD=73) due to changes in the diagnosis. All demographical, clinical, and imaging data were collected at two time points one year apart. The diagnostic criteria for MCI patients were MMSE scores between 24-30 (inclusive), a memory complaint, having objective memory loss measured by education-adjusted scores on Wechsler Memory Scale-Revised (WMS-R) Logical Memory II, a CDR of 0.5, largely preserved activities of daily living, and an absence of dementia. AD patients were diagnosed using the following criteria: MMSE scores between 20-26 (inclusive), CDR of 0.5 or 1.0, and meeting the National Institute of Neurological and Communicative Disorders and Stroke–Alzheimer’s Disease and Related Disorders Association (NINCDS-ADRDA) criteria for probable AD.

### 2.3. Cognitive assessments

The Mini-Mental State Exam (MMSE), Montreal Cognitive Assessment (MoCA), Clinical Dementia Rating scale Sum of Boxes scores (CDR-SB) and Alzheimer’s Disease Assessment Scale-Cognitive (ADAS-COG) were extracted from ADNI to evaluate cognitive function of subjects. The MMSE and MoCA are rated based on a 30-point scale. The MMSE was developed in 1975 as a simplified battery to examine cognitive status with 11 questions [12,13]. The MoCA, similar to the MMSE, is a brief but newer battery devised in 2005 [14]. The CDR-SB measures both cognitive and functional impairment at the same time. The test has many advantages over CDR global score: 1-being easier to calculate, 2-can be treated as interval data in statistical analysis, and 3-being more accurate for tracking cognitive changes across time [15]. The ADAS-11 is a brief cognitive test battery that assesses learning, memory, language production, language comprehension, constructional praxis, ideational praxis, and orientation. It includes both subject-completed tests and observer-based assessments [16,17]. The ADAS-13, the extended version of ADAS-11, evaluates two further tasks, namely a test of delayed word recall and a number cancellation or maze task [17,18].

### 2.4. Plasma NfL measurement in ADNI

The Single Molecule array (Simoa) technique was used to analyze plasma NfL levels. The combination of monoclonal antibodies and purified bovine NfL is considered a calibrator in this technique. All samples were measured twice, except for one (due to technical reasons). Analytical sensitivity was lesser than 1.0 pg/ml. In addition, there were no samples containing NfL levels in plasma below the limit of detection (LOD). Plasma samples were collected after fasting overnight (minimum 6 hours) using a 10mL lavender top tube which was gently mixed by inversion 10-12 times. The collection tubes were centrifuged at room temperature within one hour of collection and then spined for 15 minutes using the Sorvall T 6000D Centrifuge (rotor H-1000B swinging bucket rotor) at 3000 rpm (1500 rcf) with the brake on. Transferring plasma from each of the two lavender top tubes to the study specific 13 mL plastic transfer tubes were done using a sterile pipette and were firmly caped with the lavender screw cap. Then, the lavender screw-capped tubes were placed in dry ice upright in a freezer.

### 2.5. APOE genotyping

APOE genotyping was carried out at screening visits for each subject. Genotyping for all samples was conducted at the National Cell Repository for AD (NCRAD). APOE e4 positive cases were defined as subjects carrying either one or two alleles.

### 2.6. Tensor-Based Morphometry calculation in ADNI

#### 2.6.1. Image Acquisition

ADNI database (http://adni.loni.ucla.edu/) was used to extract preprocessed MR scans results (1.5T or 3T). The standard Mayo Clinic processing pipeline had been used to process images.

#### 2.6.2. Image pre-processing

For linear registration, a 9-parameter registration method was applied to match a follow-up scan to its corresponding screening scan. This leads to adjustment for linear drifts in head position and scale within the same subject. Global differences in brain scale through subjects were accounted by registering the mutually aligned time series of scans to the International Consortium for Brain Mapping template (ICBM-53). To remove one source of bias in analyzing longitudinal data, both screening and follow-up scans were resampled once during the linear registration. Then, a minimal deformation target (MDT) was constructed in 4 steps to serve as an unbiased average template image using the scans of 40 randomly selected normal subjects [19].

#### 2.6.3. Cross-sectional TBM

3D patterns of volumetric brain differences were quantified by aligning all individual screening images (N=817) to the MDT using a non-linear inverse consistent elastic intensity-based registration algorithm. This process resulted in the gradients of the deformation field which subsequently was used to elicit a Jacobian matrix field or Jacobian maps. The determinant of the local Jacobian matrix represents local volume differences in the temporal lobe of the brain. Determinants greater or lesser than one illustrates expansion or contraction relative to the normal group template, respectively.

#### 2.6.4. Longitudinal TBM

In this part, a step was added to create brain masks and remove the image background before longitudinal linear registration. This step called skull-stripping improved the precision of longitudinal TBM since subtle changes were considered as brain degeneration over time. Individual Jacobian maps were elicited by wrapping the 9P-registered and skull-stripped follow-up scan to match the corresponding screening scan using a non-linear inverse consistent elastic intensity-based registration algorithm.

#### 2.6.5. Numerical summaries of cumulative temporal lobe atrophy

A single-numerical summary of the 3D map of brain atrophy for each subject was computed to estimate the amount of cumulative temporal lobe atrophy in statistical or anatomical region of interest (ROI) in the temporal lobe of the brain. The numerical summary was derived by taking an average in a ROI. Lower TBM index shows greater degree of atrophy.

In this paper, we downloaded statistical-ROI from ADNI which was defined based on voxels with significant atrophic rates over time (p<0.0001) within the temporal lobes using a training set of 20 AD patients scanned at baseline and 12-month. For a detailed explanation on TBM processing, download the provided documentation in http://adni.loni.ucla.edu/.

### 2.7. Statistical analysis

The Statistical Package for Social Sciences (SPSS) 25 software was used for data analysis. Chi-score and Kruskal-Wallis test was performed to compare qualitative and quantitative variables, respectively. In addition, to compare longitudinal TBM and plasma NfL, Wilcoxon signed rank test was applied. Linear regression modeling was used to determine the association between TBM and plasma NfL and cognitive scores (MMSE, MoCA, CDR-SB, ADAS-11, ADAS-13) in different diagnostic groups. Sex, age, education years, and APOE e4 status were considered as covariates in the regression model. To address multiple comparison issues, the Bonferroni correction was applied. The statistical significance level was set at <0.05.

## 3. Results

### 3.1. Patient characteristics

A total of 480 participants were enrolled. The demographic and clinical characteristics of subjects at baseline and first year are shown in Table 1 and supplementary material, respectively. The male to female ratio, age, and education years at baseline were not significantly different among subgroups. Regarding baseline biomarkers, MoCA and MMSE scores, unlike CDR-SB scores, were significantly lower in AD. Plasma NfL levels varied significantly across subgroups, with the highest NfL levels in AD. Our TBM index also varied significantly, being lowest in AD.

**Table 1.** Demographic and clinical characteristics of subjects at baseline.

| Variable | CN<br>(N=157) | MCI<br>(N=278) | AD<br>(N=45) | *p-value* |
| --- | --- | --- | --- | --- |
| **Sex (Female/Male)** | 77/80 | 123/155 | 18/27 | 0.468 |
| **Age (years)**<br>**Mean (SD)** | 72.43 (6.48) | 71.38 (7.35) | 73.91 (7.49) | 0.060 |
| **Education (years)**<br>**Mean (SD)** | 16.76 (2.48) | 16.32 (2.64) | 15.93 (2.32) | 0.091 |
| **MOCA**<br>**Mean (SD)** | 26.10 (2.48) | 23.94 (2.99) | 19.07 (4.78) | <0.001*<sup>a</sup> |
| **MMSE**<br>**Mean (SD)** | 28.83 (1.23) | 27.86 (1.97) | 23.44 (3.86) | <0.001*<sup>a</sup> |
| **CDR-SB**<br>**Mean (SD)** | 0.12 (0.29) | 1.33 (1.06) | 4.38 (2.34) | <0.001*<sup>a</sup> |
| **ADAS-11**<br>**Mean (SD)** | 5.11 (2.81) | 8.30 (4.52) | 18.15 (6.95) | <0.001*<sup>a</sup> |
| **ADAS-13**<br>**Mean (SD)** | 7.81 (4.16) | 13.14 (6.87) | 27.89 (8.63) | <0.001*<sup>a</sup> |
| **APOE e4 (Yes/No)** | 44/113 | 122/156 | 37/8 | <0.001* |
| **Plasma NFL (pg/ml)**<br>**Mean (SD)** | 35.63 (16.79) | 38.28 (23.86) | 52.12 (27.24) | <0.001*<sup>b</sup> |
| **TBM**<br>**Mean (SD)** | 992.71 (7.96) | 989.43 (9.92) | 979.04 (13.34) | <0.001*<sup>a</sup> |

*Note:*
Significant level is <0.05, Chi-score test and Kruskal-Wallis test were used
Multiple comparison p-values were corrected by Bonferroni method;
a: CN>MCI, CN>AD, MCI>AD
b: CN<AD, MCI<AD, MCI=AD

### 3.2. Correlations between plasma NfL and TBM index

We assessed the correlation between the TBM index and plasma NfL levels at baseline and one-year follow-up (Figure 1). A significant association between TBM index and plasma NfL was found only in the pooled subject analysis (β=-0.139, *P*=0.004). Subgroup analyses revealed no significant associations. One-year follow-up showed no significant correlations.

**Figure 1.**
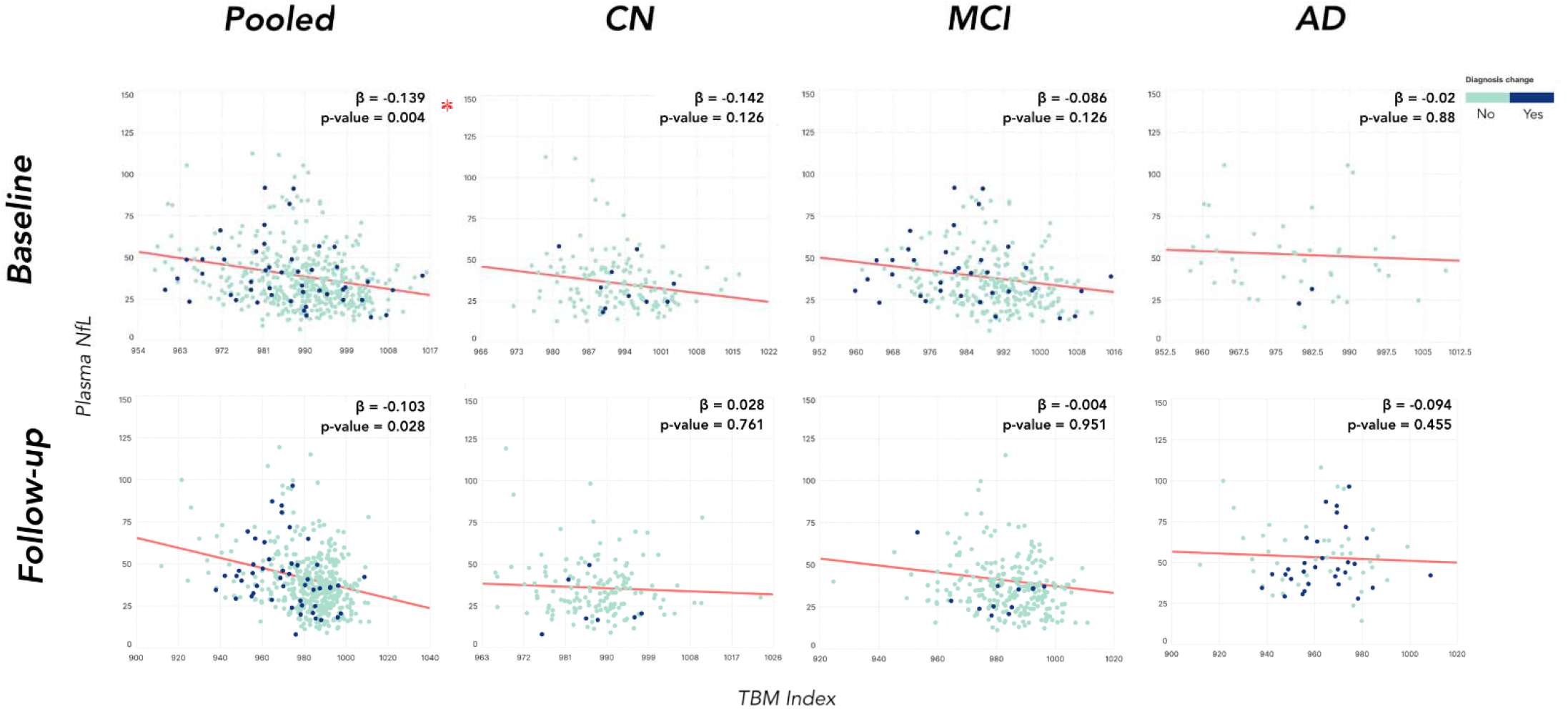
Associations between baseline and follow up plasma NfL measures with TBM measure of the temporal lobe region of interest Significant level is <0.005, Correlation was adjusted by sex, age, APOE e4, and education, Asterisk shows significant p-values also after correction

### 3.3. Longitudinal comparisons of Plasma NfL levels and TBM index

Table 2 shows p-values of paired comparisons. Of the 480 participants, 147, 241, and 43 remained unchanged diagnosis as CN, MCI, and AD, respectively. Notably, 30 individuals converted from MCI to AD (MCI-AD group). Plasma NfL levels increased significantly in MCI after one year (*P*<0.001). TBM index decreased significantly in all groups (*P*<0.001 for all groups), indicating progression of temporal lobe atrophy. Notably, a significant increase in plasma NfL (*P*<0.001) coupled with a significant decrease in TBM measurements (*P*<0.001) was only observed in the MCI group.

**Table 2.** p-values of two time-point comparisons of Plasma NFL and TBM.

| **Table 2 p-values of two time-point comparisons of Plasma NFL and TBM** |  |  |  |  |  |  |  |
| --- | --- | --- | --- | --- | --- | --- | --- |
| Variable | CN – CN<br>(N=147) | MCI – MCI<br>(N=241) | AD – AD<br>(N=43) | MCI – AD<br>(N=30) | MCI -CN<br>(N=7) | CN-MCI<br>(N=10) | AD-MCI<br>(N=2) |
| **Plasma NFL** | 0.612 | <0.001* | 0.031<sup>a</sup> | 0.162 | NA | NA | NA |
| **TBM** | <0.001* | <0.001* | <0.001* | <0.001* | NA | NA | NA |

*Note:*
Significant level is <0.05, Wilcoxon Signed Ranks Test was used, a: not significant after correction, NA=not applicable due to very small sample size, Asterisk shows significant p-values also after correction

### 3.4. Comparing the average rate of change of TBM and plasma NfL in different diagnostic groups over one year

Table 3 highlights the average rate of change in TBM and Plasma NfL over one year. The AD group showed the greatest average decrease in TBM, followed by MCI-AD, MCI, and CN groups. A significant difference in TBM average change rate was observed between subgroups (*P*<0.001; MCI-AD>CN, MCI-AD>MCI, AD>CN, AD>MCI, MCI-AD=AD-AD, MCI=CN). Regarding plasma NfL, the MCI group showed the largest average increase. Statistically significant difference in plasma NfL average change rate was only observed between CN and AD group (*P*=0.013).

**Table 3.** The rate of change of TBM and Plasma NfL between baseline and year one.

|  |  |  |  |  |  |
| --- | --- | --- | --- | --- | --- |
| Variable | CN-CN<br>(N=147) | MCI-MCI<br>(N=241) | AD-AD<br>(N=43) | MCI-AD<br>(N=30) | p-value |
| TBM rate<br>Mean (SD) | -4.61 (9.01) | -6.24 (9.13) | -19.14 (12.34) | -16.35 (8.06) | <0.001*<sup>a</sup> |
| Plasma NfL rate<br>Mean (SD) | 0.61 (13.88) | 3.75 (32.36) | 1.87 (16.06) | 1.03 (23.70) | 0.013*<sup>b</sup> |

*Note:*
Significant level is <0.05, Kruskal-Wallis test was used
Multiple comparison p-values were corrected by Bonferroni method;
a: MCI-AD>CN-CN, MCI-AD>MCI-MCI, AD-AD>CN-CN, AD-AD>MCI-MCI, AD-AD=MCI-AD, MCI-MCI=CN-CN
b: AD-AD>CN-CN, all other comparisons showed non-significant p-values

### 3.5. Correlations between plasma NfL and TBM index with cognitive scores

Table 4 presents correlations between TBM, plasma NfL, and cognitive scores at baseline. In MCI participants, TBM showed significant correlations with all cognitive scores (MMSE, MoCA, CDR-SB, ADAS-11, ADAS-13), unlike plasma NfL. In AD patients, TBM correlated significantly with all cognitive scores except for CDR-SB, while plasma NfL did not show significant correlation with all cognitive scores.

**Table 4.**
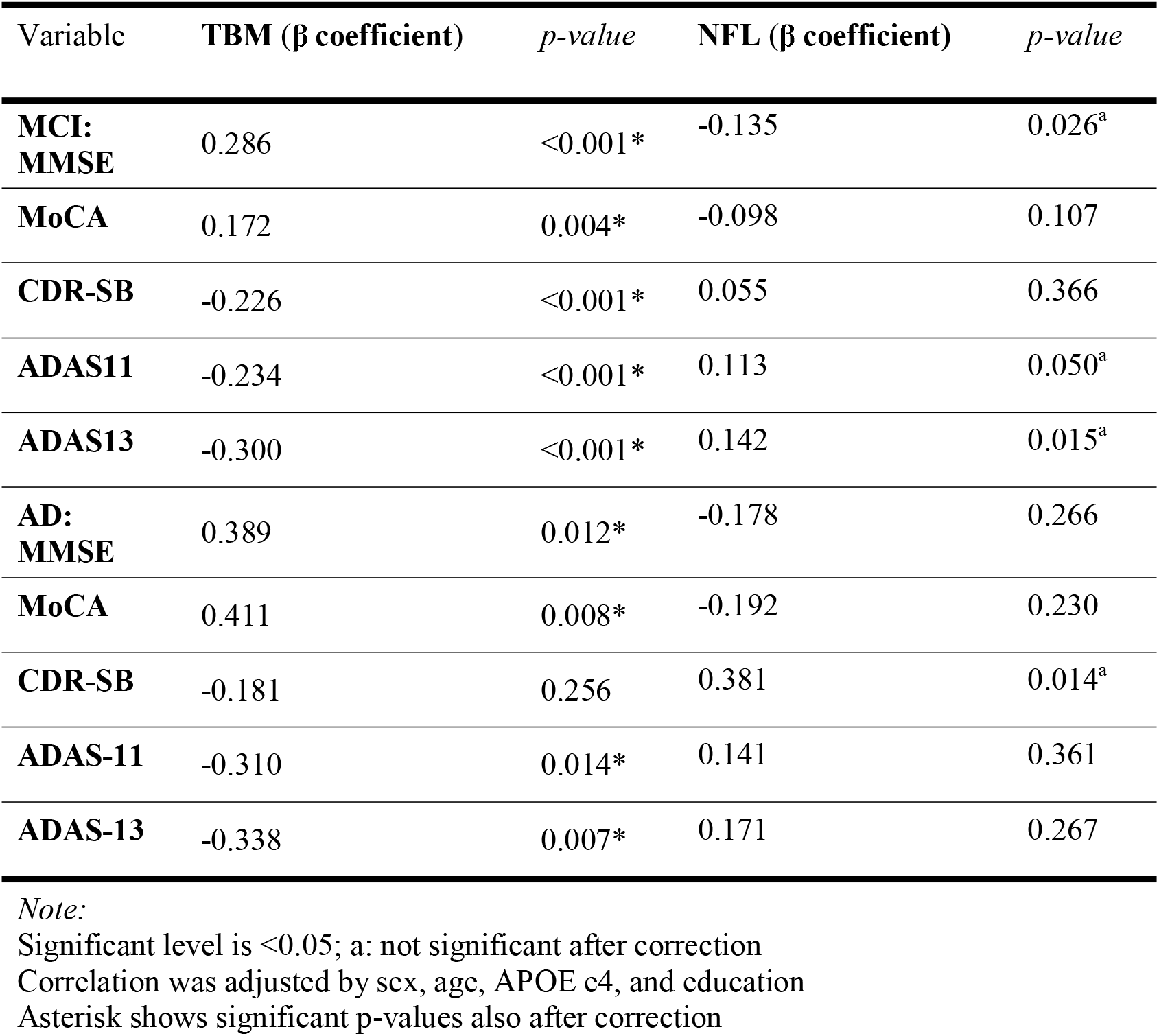
TBM and plasma NFL in correlation with cognitive scores at baseline.

|  |  |  |  |  |
| --- | --- | --- | --- | --- |
| Variable | TBM ($\beta$ coefficient) | *p*-value | NFL ($\beta$ coefficient) | *p*-value |
| **MCI:**<br>**MMSE** | 0.286 | <0.001* | -0.135 | 0.026<sup>a</sup> |
| **MoCA** | 0.172 | 0.004* | -0.098 | 0.107 |
| **CDR-SB** | -0.226 | <0.001* | 0.055 | 0.366 |
| **ADAS11** | -0.234 | <0.001* | 0.113 | 0.050<sup>a</sup> |
| **ADAS13** | -0.300 | <0.001* | 0.142 | 0.015<sup>a</sup> |
| **AD:**<br>**MMSE** | 0.389 | 0.012* | -0.178 | 0.266 |
| **MoCA** | 0.411 | 0.008* | -0.192 | 0.230 |
| **CDR-SB** | -0.181 | 0.256 | 0.381 | 0.014<sup>a</sup> |
| **ADAS-11** | -0.310 | 0.014* | 0.141 | 0.361 |
| **ADAS-13** | -0.338 | 0.007* | 0.171 | 0.267 |

*Note:*
Significant level is <0.05; a: not significant after correction
Correlation was adjusted by sex, age, APOE e4, and education
Asterisk shows significant p-values also after correction

Similar results as baseline visit were also replicated for the first-year visit (shown in Table 5). Note that inclusion of CN subjects in the analysis did not provide statistically significant p-values, and thus, they were reported in the supplementary materials.

**Table 5.** TBM and plasma NFL in correlation with cognitive scores at first year.

| Variable | TBM ($\beta$ coefficient) | *p*-value | NFL ($\beta$ coefficient) | *p*-value |
| --- | --- | --- | --- | --- |
| **MCI:**<br>**MMSE** | 0.228 | <0.001* | 0.041 | 0.518 |
| **MoCA** | 0.241 | <0.001* | -0.042 | 0.507 |
| **CDR-SB** | -0.243 | <0.001* | -0.004 | 0.947 |
| **ADAS-11** | -0.363 | <0.001* | 0.009 | 0.889 |
| **ADAS-13** | -0.388 | <0.001* | 0.030 | 0.637 |
| **AD:**<br>**MMSE** | 0.552 | <0.001* | -0.060 | 0.622 |
| **MoCA** | 0.457 | <0.001* | -0.129 | 0.292 |
| **CDR-SB** | -0.281 | 0.019<sup>a</sup> | 0.300 | 0.010<sup>a</sup> |
| **ADAS11** | -0.384 | <0.001* | 0.155 | 0.162 |
| **ADAS13** | -0.401 | <0.001* | 0.117 | 0.291 |

*Note:*
Significant level is <0.05; a: not significant after correction
Correlation was adjusted by sex, age, APOE e4, and education
Asterisk shows significant p-values also after correction

## 4. Discussion

Our study investigated the relationship between the TBM as a measure of temporal lobe atrophy and the plasma NfL as a measure of axonal degeneration in Alzheimer’s dementia spectrum at baseline and one-year follow-up. Plasma NfL levels and TBM indices were significantly higher at more advanced stages of dementia (AD>MCI>CN). We found a significant but very weak association between our TBM index and plasma NfL only when all subjects were pooled together, and no significant associations were found between either biomarker in CN, MCI, or AD subjects. In our longitudinal analyses, we found a consistent pattern of significant temporal lobe atrophy in all groups over time, while plasma NfL levels increased insignificantly. Finally, significant increases in plasma NfL concentrations were coupled with significant decreases in TBM measurements after a one-year follow-up only in MCI group.

Our findings of temporal lobe changes in AD dementia are in line with a long history of such findings. Previous studies report synapse loss [20], volume changes [21], and neurofibrillary tangle formation [22] in the temporal lobe during aging and dementia. These changes were captured using TBM, where changes were able to predict disease progression in all stages, even in healthy controls [11]. Measures of temporal atrophy derived from this method have been shown to correlate with decline in MMSE, CDR, and logical/verbal learning memory scores, as well as higher CSF p-tau levels in the AD continuum [23]. In line with these findings, we observed a significant decrease in the TBM index in patients that progressed from MCI to AD during one-year follow-up. Additionally, the rate of change of temporal lobe atrophy was significantly higher in patients with MCI who progressed to AD, compared to those who did not.

NfL chain levels in serum are a marker of neurodegeneration in Alzheimer’s disease (AD). Previous research indicates that NfL is a non-specific marker of neurodegeneration, with increased levels in CSF observed in various dementias[24]. Studies show that elevated levels of NfL in CSF and plasma are linked to AD-related neuroimaging findings, including hippocampal atrophy, ventricular enlargement, and cortical thinning [25–27]. Serum NfL levels also increase in healthy aging people and predict brain volume loss [28]. A longitudinal study found that increased NfL levels correlate with cognitive deficits and several markers for AD [29]. The rate of change of NfL levels is also informative about the disease process, with accelerated increases correlating with more rapid cognitive decline, similar to Preische et al. [30]. However, our analysis of plasma NfL did not replicate this finding. Additionally, comparing these results with the abovementioned association between the rate of temporal atrophy and disease progression is another example of the superiority of TBM index as a biomarker in Alzheimer’s dementia. Indeed, our results show that TBM measures are more sensitive than plasma NfL levels to longitudinal brain alterations: two time-point comparisons and average rates of change of our TBM index are significantly different in all subgroups while plasma NfL levels are not. Accordingly, we might expect TBM to be a more robust tool in the following-up of dementia spectrum than plasma NfL. Our findings regarding the association of these biomarkers with cognitive scores support this conclusion as well. In particular, TBM values were significantly associated with all investigated cognitive scores in the MCI subgroup and MMSE, MoCA, ADAS-11 and ADAS-13 scores in the AD subgroup. On the other hand, NfL levels were not associated with all cognitive scores in both MCI and AD groups. Plasma NfL’s inability to predict any cognitive score in MCI group further points to the unsuitability of this marker in the early stages of the disease. These results align with a community-based study by Carmen Lage-Martinez et al., which found that plasma NfL was not effective in detecting preclinical AD [31]. However, a study by Hao Hu et al. showed that plasma NfL was valuable for evaluating neurodegeneration and predicting disease progression in individuals with preclinical AD [32]. Our results concerning the utility of TBM in early dementia diagnosis and cognitive score prediction support the potential of TBM-based analysis to monitor structural atrophy in AD during its early stages, before severe cognitive impairment occurs [10,11]. Finally, it must be restated that despite the inability of plasma NfL in predicting the dementia progression, previous results show that it might be able to differentiate between CN, MCI and AD.

Our study provides further evidence for the utility of TBM and serum NfL as imaging and fluid biomarkers of brain health. We observed a significantly increased level of plasma NfL in the MCI groups at one-year follow-up compared to baseline but a significant reduction in TBM measurements in all groups, indicating that our TBM index might be a more sensitive marker of cortical atrophy than NfL. Contrary to our expectations, the group with the highest rate of TBM change did not show a similarly high rate of NfL change. Importantly, no significant increase in NfL levels were observed in the subgroup that progressed from MCI to AD in our follow-up, showcasing the limitations of plasma NfL levels as a predictor of dementia progression. These results must however be interpreted with caution, given the small size of the MCI-AD subgroup (N=30). Furthermore, we observed a significant association between the two variables only in the pooled analysis, suggesting that plasma NfL levels might change at a different rate compared to the TBM index leading to a poor correlation. Overall, while temporal lobe atrophy and axonal degeneration may occur together in AD, this co-occurrence does not manifest itself clearly in measurements from TBM and plasma NfL; in other words, lower measured TBM indices are not robustly correlated with higher plasma NfL levels.

To the best of our knowledge, this is the first study to investigate the relationship of plasma NfL and temporal lobe atrophy using TBM in the Alzheimer’s dementia spectrum.

The results of this study should be interpreted within the context of several limitations. The TBM index we used in our study only measured temporal lobe atrophy, and did not consider other cortical or subcortical areas or any other measures of brain health. Finally, TBM requires significant computational resources, including high-performance computing infrastructure and specialized software. The processing time and resources required can be a limiting factor for researchers with limited access to these resources.

Another important limitation of this study is the lack of subgroup analyses of less common forms of AD, such as early-onset and familial forms. These subtypes may have different underlying mechanisms, disease progression rates, and responses to treatment. Consequently, certain biomarkers, may be more pronounced in specific subtypes [33].

## 5. Conclusion

In conclusion, our study provides further evidence for the utility of TBM and serum NfL as imaging and fluid biomarkers of brain health. our findings indicate that plasma NfL is not reliably associated with TBM, and is less effective and sensitive than TBM in predicting dementia progression and cognitive performance. Further studies in larger cohorts of subjects with different age groups and with different imaging techniques and biomarkers are warranted in order to confirm these findings and to better elucidate the relationship between these biomarkers and current diagnostic tools.

## Author contributions

Nikan Amirkhani: data interpretation, writing original draft

Amirhossein Khodadadi: study design, data collection, data analysis, data interpretation, writing original draft

Zahra Nouri: data collection, writing original draft

Tanin Adlparvar: data collection, writing original draft

Sanaz Eskandari: writing original draft

Rezvan Barzgar: writing original draft

Pouya Sojoudi: writing original draft

Mahsa Mayeli: developing the research idea, data collection, interpreting the results, supervising the research and revising the manuscript

## Data Availability

All data produced in the present study are available upon applying for ADNI database.

https://adni.loni.usc.edu/

## Acknowledgements

We would like to acknowledge the Alzheimer’s Disease Neuroimaging Initiative (ADNI) for providing the data of this paper.

## Supplementary materials

Supplementary material associated with this article can be found, in the online version.

