## supplementary material for "Axonal degeneration serum markers and temporal lobe atrophy in Alzheimer’s dementia continuum: a longitudinal study of plasma neurofilament light and tensor-based morphometry"

| **Table 1 Demographic and clinical c*haracteristics of subjects at first year*** | | | | |
| --- | --- | --- | --- | --- |
| Variable | CN  (N=154) | MCI  (N=253) | AD  (N=73) | *p-value* |
| **Sex (Female/Male)** | 73/81 | 113/140 | 32/41 | 0.828 |
| **Age (years)**  **Mean (SD)** | 72.23 (6.44) | 71.35 (7.38) | 73.48 (7.40) | 0.037* |
| **Education (years)**  **Mean (SD)** | 16.90 (2.47) | 16.28 (2.64) | 15.95 (2.41) | 0.016* |
| **MOCA**  **Mean (SD)** | 26.18 (2.46) | 24.26 (3.17) | 16.93 (4.80) | <0.001*^a^ |
| **MMSE**  **Mean (SD)** | 29.08 (1.14) | 27.90 (2.10) | 22.23 (4.03) | <0.001*^a^ |
| **CDR-SB**  **Mean (SD)** | 0.09 (0.28) | 1.28 (1.01) | 5.25 (2.40) | <0.001*^a^ |
| **ADAS 11**  **Mean (SD)** | 4.91 (2.88) | 8.20 (4.70) | 20.30 (8.81) | <0.001*^a^ |
| **ADAS 13**  **Mean (SD)** | 7.71 (4.16) | 13.11 (7.16) | 30.56 (10.63) | <0.001*^a^ |
| **APOE e4 (Yes/No)** | 42/112 | 108/145 | 53/20 | <0.001* |
| **Plasma NFL (pg/ml)**  **Mean (SD)** | 35.80 (18.93) | 40.62 (33.28) | 53.24 (19.44) | <0.001*^b^ |
| **TBM**  **Mean (SD)** | 987.99 (9.09) | 983.93 (12.57) | 961.68 (18.34) | <0.001*^a^ |
| *Note:*  Significant level is <0.05  Chi-score test and Kruskal-Wallis test were used  Multiple comparison p-values were corrected by Bonferroni method  Asterisk shows significant p-values after correction  a: CN>MCI, CN>AD, MCI>AD, b: CN<AD, MCI<AD | | | | |

| **Table 2** | | | | |
| --- | --- | --- | --- | --- |
| Variable | **TBM** (**β coefficient**) | *p-value* | **NFL** (**β coefficient)** | *p-value* |
| **Baseline:**  **MMSE** | -0.062 | 0.443 | 0.140 | 0.085 |
| **MoCA** | -0.065 | 0.426 | -0.081 | 0.319 |
| **CDR-SB** | 0.113 | 0.163 | 0.011 | 0.889 |
| **ADAS-11** | -0.097 | 0.255 | 0.008 | 0.916 |
| **ADAS-13** | -0.128 | 0.152 | 0.008 | 0.919 |
| **First year:**  **MMSE** | 0.032 | 0.700 | 0.087 | 0.291 |
| **MoCA** | 0.101 | 0.219 | -0.138 | 0.091 |
| **CDR-SB** | 0.013 | 0.875 | 0.001 | 0.988 |
| **ADAS-11** | -0.135 | 0.115 | 0.108 | 0.162 |
| **ADAS-13** | -0.158 | 0.068 | 0.088 | 0.261 |
| *Note:*  Significant level is <0.05; correlation was adjusted by sex, age, APOE4, and education | | | | |

Table 2 provides the reader with the results of correlation between cognitive scores and biomarkers in cognitively normal subjects at baseline and first-year, respectively.
